# Adaptation and Psychometric Validation of a Facility-Level Tool to Assess Telemedicine Readiness in Primary Care

**DOI:** 10.64898/2026.07.01.26356790

**Authors:** Stefan Escobar-Agreda, David Villareal-Zegarra, C. Mahony Reategui-Rivera, Yscenia Paredes-Gonzales, Leonardo Rojas-Mezarina

**Affiliations:** Escuela de Posgrado, Universidad San Ignacio de Loyola, Lima, Peru; IDEAS Research Group, School of Medicine, Universidad Científica del Sur, Lima, Peru; Digital Health Research Center, Digital Health S.A., Lima, Peru; Instituto Peruano de Orientación Psicológica, Lima, Peru; Unidad de Telesalud, Facultad de Medicina, Universidad Nacional Mayor de San Marcos, Lima, Peru

**Author notes:** Corresponding author: David Villareal-Zegarra.

**Keywords:** telemedicine, telehealth, readiness, psychometric validation, health facilities, primary care, organizational readiness, maturity model, measurement invariance, structural validity, criterion validity, implementation sciences

## Abstract

**Background:** Telehealth has expanded rapidly, yet its sustained use beyond emergency responses remains uneven. Facility and organizational conditions are modifiable determinants of healthcare intervention implementation, and readiness assessments are key steps in the early adoption and further uptake process. However, many telemedicine readiness assessment instruments lack psychometric evidence, limiting their value for benchmarking, prioritizing investments, and monitoring progress at scale.

**Objective:** To adapt and psychometrically validate the Telemedicine Readiness Inventory at the Facility Level (TRI-F).

**Methods:** We conducted a cross-sectional analysis using data from an online assessment of a large sample of primary care facilities (PCFs) in Peru, held between December 2023 and March 2024. The analytic sample included 774 PCFs, with one designated respondent per facility completing the survey. Internal structure was evaluated using exploratory factor analysis and confirmatory factor analysis. Internal consistency was estimated using Cronbach’s alpha and McDonald’s omega. Measurement invariance was evaluated across the facility complexity and the respondant’ time working at the PCF. Criterion validity was examined using Spearman correlations between the five domain scores (Organizational readiness, Processes, Digital environment, Human resources, and Regulatory issues) and proportion of telemedicine modality among overall outpatient encounters.

**Results:** Confirmatory factor models showed adequate fit across domains, with CFI values ranging from 0.959 to 0.999, TLI from 0.952 to 0.997, RMSEA from 0.028 to 0.065, and SRMR from 0.016 to 0.057, for the five domains assessed. Internal consistency was acceptable to high across all domains (α = 0.75-0.87; ω = 0.76-0.88). Measurement invariance was supported across the facility category and time working at the PCF, with ΔCFI values below 0.010. Criterion validity analyses showed positive but small correlations between all five domains and proportional telemedicine use (r_s_= 0.15–0.23; p < 0.001).

**Conclusions:** The adapted tool showed satisfactory structural validity, internal consistency, and measurement invariance for assessing telemedicine readiness in PCFs. The availability of percentile-based norms supports interpretation and benchmarking. The instrument can support implementation planning, monitoring, and prioritization of technical assistance in primary care settings.

## Introduction

Telemedicine is increasingly viewed as a core strategy to expand access and continuity of healthcare, particularly in primary care settings, where geographic barriers and workforce constraints can limit timely service delivery [1–3]. During the COVID-19 pandemic, telemedicine expanded rapidly as health systems adapted to infection control measures and restrictions on face-to-face care [1,4]. However, reviews in low-and-middle-income countries (LMICs) noted that this scale-up often faced challenges for long-term continuity, especially in settings with persistent infrastructural and regulatory constraints [5,6]. In Peru, an upper-middle-income country, telemedicine appointments increased during the pandemic but subsequently declined, reflecting the challenge of consolidating telemedicine as routine care rather than a temporary response [7].

Sustained telemedicine implementation in LMICs rely on multiple determinants operating at the patient, provider, organizational and healthcare-system levels [8]. Patient-side barriers, particularly limited access to connectivity and appropriate devices, and variable digital literacy; which can make telemedicine acceptability and effective use uneven across populations [6,9]. However, many of these constraints are difficult to be addressed solely by the health sector, as those are rooted in broader structural barriers (e.g. telecommunication infrastructure, nationwide educational programs, affordability and budget constraints) and cross-sectoral policies [10,11]. In this context, telemedicine uptake also depends heavily on healthcare provider and organizational adoption, because those determine whether these services are implemented and embedded into routine workflows in ways that support continued everyday use [12,13]. From a managerial and policy perspective, provider and organizational readiness is an efficient measure because it can be influenced through organizational and change management strategies, including leadership and governance support, resourcing, workforce training, workflow integration, and operational support [14,15].

Organizational readiness for change refers to the shared commitment and perceived collective capability of organization’ members to implement a change [16]. When readiness is high, organizations are more likely to initiate, routinize and sustain new interventions [17]. Additionally, readiness was reported to be associated with higher intervention implementation fidelity [18–20]. In telemedicine, readiness has been found as multidimensional, commonly encompassing human resources, technical infrastructure, workflows/processes, governance, and policies [21]. Readiness assessments could therefore help decision-makers identify modifiable gaps before scale-up and monitor whether health facilities are prepared to sustain telemedicine services [22,23]. However, these assessments must produce reliable, comparable and valid scores in the specific contexts where they are used [24].

Multiple frameworks and instruments have been proposed to assess whether health facilities met the conditions required to implement and sustain telemedicine [25]. However, their psychometric evaluation is often sparse, limiting their use as reliable quantitative tools for comparisons across settings and over time [26]. During the COVID-19 pandemic, the Pan American Health Organization (PAHO) and the Inter-American Development Bank (IDB) released a tool to assess health institutions’ readiness to implement telemedicine services [27], but its psychometric performance remains unknown. In Peru, this tool was modified and used for the assessment of primary care facilities (PCFs). This study evaluated the psychometric performance of the adapted tool among a nationwide survey among PCFs in Peru, including its internal structure, reliability, and criterion-related validity. As criterion-related validity evidence, we examined whether readiness scores were associated with facility-level telemedicine adoption.

## Methods

### Design

Observational, cross-sectional psychometric validation study following COSMIN standards for structural validity, content validity, and internal consistency. Our study is a secondary data analysis of a telemedicine evaluation conducted by the Peruvian Ministry of Health’s Directorate of Telehealth. The data are publicly accessible and were obtained through a formal request.

### Setting

The Peruvian health system is fragmented and decentralized, with the Ministry of Health (MoH) as the main public provider of primary care services and a central point of access for institutional care in the public sector. In 2025, among people with a health problem who sought care in healthcare facilities, 58.3% attended MoH facilities showing this health subsystem concentrates the largest share of healthcare demand in Peru [28]. Peru has also developed a regulatory and policy framework to support telehealth implementation. The Telehealth Law was enacted in 2016, and the National Telehealth Plan 2020–2023 established strategic actions to strengthen telehealth services, particularly in rural areas and settings with limited health service capacity [29,30]. These actions included improving technological infrastructure, connectivity, interoperability, biomedical and information technology equipment, logistical support, and workforce capacity for telehealth implementation.

In this context, the MoH made substantial investments to strengthen the National Telehealth Network in primary care facilities. For example, the MoH reported an investment of 53,152,974.71 PEN for the acquisition of 16,954 information technology devices and accessories, as well as 3,121 biomedical devices, to be distributed across 678 primary care health facilities nationwide [31].

However, the availability of equipment and regulatory support has not necessarily translated into sustained telemedicine use. Evidence suggests that telemedicine use declined substantially after the COVID-19 pandemic and varied across regions and facility contexts, reflecting persistent differences in connectivity, local capacity, workflows, demand, and organizational readiness [32].

This implementation gap highlights the need to assess not only whether facilities have telemedicine-related equipment or formal regulatory support, but also whether they have the organizational, procedural, technological, human-resource, and regulatory conditions required to integrate telemedicine into routine care.

### Data Source

The information of the readiness tool comes from a national assessment conducted in a large sample of PCFs via a secure, self-administered online survey held by the MoH’s Directorate of Telehealth between December 2023 and March 2024. PCFs across all 25 regions of Peru that were part of the National Telehealth Network were recruited for the survey. In Peru, this Network includes MoH PCFs that have demonstrated consistent telemedicine productivity and whose inclusion has been reviewed and approved in consultation with the telehealth technical team in each region. The survey also contains information regarding the respondent and the PCF.

The metric for the criterion validity analysis was collected from the Health Information System (HIS), an anonymized administrative dataset obtained through a formal request submitted via the Peruvian transparency portal, which includes nationwide records of healthcare encounters among all the MoH healthcare facilities between January and June 2024. Additionally, the Peruvian Current Procedural Terminology (P-CPT) dataset and the 2024 National Health Facilities Repository of Peru, both publicly available through the Peruvian government open data portal, were also consulted to delimit outcome operationalization. These datasets were linked to the national readiness assessment database using the PCFs unique ID code.

### Participants

The study used data from a facility-level online survey administered to PCFs in the National Telehealth Network. Although the original MoH assessment used stratified probability sampling by region and included an initial sample of 826 PCFs, additional facilities outside the original sampling frame also completed the survey, resulting in 993 available records. Because this study focused on the psychometric performance of the adapted instrument rather than on estimating representative readiness scores, we included all facilities that completed the survey and met the eligibility criteria. Eligible responses were those completed by the facility head or the facility’s telehealth coordinator/focal point and with complete information for the variables required in the analysis.

For the present study, we estimated a minimum sample size of 274 participants for the confirmatory factor analysis. We assumed an expected CFI of 0.95, an average factor loading of 0.6, a statistical power of 80%, six items, and a significance level (α) of 0.05. We considered only the sample size for the six-item dimension, as it required the largest sample. The sample size calculation for each dimension is presented in Supplementary Material 1.

### Variables and Tool Development

#### Telemedicine Readiness Inventory at the Facility Level (TRI-F)

The assessment tool was originally developed by PAHO and IDB [27], and subsequently adapted by the technical team of the MoH Telehealth Directorate. The tool adaptation is reported here for transparency rather than as part of the present research. The original instrument comprised 85 items across six domains: Organizational Readiness, Processes, Digital Environment, Human Resources, Regulatory Issues, and Expertise. Following expert content review, only 53 items were included, and the Expertise domain was excluded because it reflects individual rather than institutional attributes, consistent with COSMIN guidance (see Table 1) [33].

**Table 1.** Domains and content areas of the MoH adapted instrument.

| Domain | No. of items | Content areas covered |
| --- | --- | --- |
| Organizational readiness | 22 | Leadership, training, infrastructure, institutional support, and telemedicine governance and monitoring mechanisms |
| Processes | 8 | Service delivery and implementation, including the existence of protocols and workflows |
| Digital environment | 12 | Connectivity, available equipment, technical support, maintenance, contingency plans, and health information systems used for documenting telemedicine services |
| Human resources | 7 | Availability and competencies of clinical personnel and information technology support staff |
| Regulatory issues | 7 | Knowledge and application of the regulatory framework, legal support, informed consent, and the facility lead's familiarity with concepts related to telemedicine implementation |

In the original tool the options of each item were on a 4-point Likert scale (1=None, 2=Beginner, 3=Advancing, and 4=Ready) and respondents had to infer how the generic labels applied to each item, which could introduce ambiguity and reduce consistency in self-administration by the designated PCF respondent. To overcome these challenge item-specific response sets were were developed so that, for each item, the four choices explicitly represented the corresponding likert levels. The MoH team iteratively refined this redesign based on feedback from key health personnel at selected PCFs. In addition, items referring to characteristics of the telemedicine information system used were removed because Peru uses a single national informatics system for telemedicine encounters, which might limited the discriminative value of those items.

Although the original instrument was framed as a maturity tool, its adapted version was used by MoH and in this study as an operational measure of facility-level telemedicine readiness. This decision was based on the content of the retained items and dimensions, which primarily assessed pre-implementation conditions relevant to telemedicine deployment, including organizational, procedural, technological, human-resource, and regulatory aspects [21]. Items reflecting prior telemedicine experience or ongoing service delivery were excluded from the readiness score. The conceptualization of readiness was informed by the telehealth readiness literature, and by prior work positioning telehealth readiness as a prerequisite for successful implementation [34,35].

#### Criterion validity indicator

Proportion of telemedicine use was defined as the percentage of telemedicine encounters as the number of telemedicine encounters divided by the total number of outpatient encounters per PCF, over the previous 6 months. This indicator was constructed by excluding all outpatient encounters whic reported health procedures according to the P-PCT, except those procedures that are performed both via telemedicine services as in-person visits (psychological interventions, counseling, orientation and education activities). This indicator is a proxy for the relative integration of telemedicine into routine healthcare delivery per PCF.

#### Co-variables

We included characteristics of the designated PCF respondent, including sex, age, profession, and tenure at the facility, as well as facility-level characteristics, including level of complexity, geographic region. Facility complexity was defined according to the scope of services and available infrastructure: facilities that mainly performed routine procedures and had limited infrastructure were classified as lower-complexity facilities, whereas those providing more complex procedures and having a larger health workforce were classified as higher-complexity facilities. Geographic area was classified using an urban-rural classification based on the Peruvian hierarchy of population centers and the national definition of rurality. We used these descriptive variables to characterize the sample and to support subgroup descriptions; we did not include them in the computation of the instrument scores. Additionally, we collected information on the responsible personnel who were evaluated, including their role within the health facility (telehealth coordinator vs. non-coordinator), and length of service (≥6 months vs. ≤5 months).

### Analysis Plan

#### Validity based on internal structure

The internal structure of the different dimensions was evaluated in two stages: an exploratory factor analysis (EFA) and a confirmatory factor analysis (CFA). Given the expected factorial complexity, we performed stratified analyses by domain. For both analyses, the sample was randomly divided into proportionally sized groups.

First, we conducted an EFA. We evaluate only one-factor models, as the instrument has already been developed and evaluated by a group of experts [27]. Only items with factor loadings greater than 0.40 were included in the analysis. We used polychoric correlation matrices to account for the ordinal nature of the variables, the weighted least squares estimator, and oblimin rotation [36].

Second, CFA was used to test the structures identified in the exploratory phase. We used the weighted least squares estimator with mean and variance adjustment and polychoric correlations. Model fit was evaluated using a set of fit indices: CFI (Comparative Fit Index) and TLI (Tucker-Lewis Index), with acceptable values defined as greater than 0.90; and SRMR (Standardized Root Mean Square Residual) and RMSEA (Root Mean Square Error of Approximation) with 90% confidence interval, with acceptable values defined as below 0.08. Correlated errors were only examined when model fit indices indicated poor fit [36,37]. Items with the highest correlation error were removed until a factor model with optimal fit indices was obtained.

#### Validity based on measurement invariance

Multiple measurement invariance models were evaluated using multigroup confirmatory factor analysis across relevant variables (i.e., facility complexity, and time working at the PCF). For each variable, hierarchical models with progressively increasing constraints were compared across categories [38]. The primary criterion for model comparison was the change in the Comparative Fit Index (ΔCFI), with ΔCFI < .01 between consecutively more constrained models indicating evidence of measurement invariance [38]. Configural, thresholds, and metric invariance were sequentially tested. ΔCFI was preferred over the χ² test due to the latter’s sensitivity to large sample sizes [38].

#### Internal consistency

Internal consistency reliability was assessed using the ordinal α (alpha) and ordinal ω (omega) coefficients. Values above 0.70 were considered acceptable [39].

#### Normative values

To support local interpretation and facilitate the use of the tool, we calculated percentile values for each dimension. Percentiles indicate the relative position of a score within the observed distribution and allow users to compare a facility score with those obtained in the study sample. We classified scores into three interpretive levels: low, from the minimum value to the 25th percentile; medium, from the 26th to the 74th percentile; and high, from the 75th percentile to the maximum value. We also reported percentile values from the 1st to the 99th percentile to provide a more detailed description of score distribution. In addition, we calculated descriptive statistics, including the mean and standard deviation. This information allows to understand how scores are distributed across each dimension and supports the interpretation of results in local digital health and biomedical informatics contexts.

#### Criterion validation

Criterion validity was examined by assessing the association between the five domains of the TRI-F and the proportion of telemedicine use. This indicator was selected as proxy of a PCFs’ operational capacity to deliver telemedicine services. Given it was numerical proportion-based variables, Spearman’s rank correlation coefficients were estimated. The magnitude of the associations was interpreted as small (r > 0.20), moderate (r > 0.50), and strong (r > 0.80).

All analyses were performed using R Studio (version 2025.05.0) with R version 4.5.1.

### Ethical aspects

This study is a secondary analysis of administrative databases and therefore poses no ethical risk to participants, as it does not include contact information or personal identifiers. Accordingly, we request an exemption from ethical review because no primary data collection was conducted. The study was submitted to the Institutional Research Ethics Committee of the Universidad Científica del Sur (CONSTANCIA N° 195-CIEI-CIENTÍFICA-2026). Our study complied with the Declaration of Helsinki guidelines.

## Results

### Description of participants

Initially, 993 records were obtained from the evaluation. After applying the inclusion criteria, 25 records were excluded because the respondents were not healthcare providers, and 13 were excluded because the facilities were not primary care but large hospitals. Subsequently, we applied the criterion of having complete data in the tool items. As a result, 181 records with missing data were excluded. Therefore, we kept data from 774 PCFs, and an inclusion rate of 77.9% of the total sample. Figure 1 presents a flow chart describing the record selection process, including the reasons for exclusion and the final number of PCFs included in the study. Table 2 presents the characteristics of the facilities included in the study and of the designated healthcare personnel who responded.

**Figure 1.**
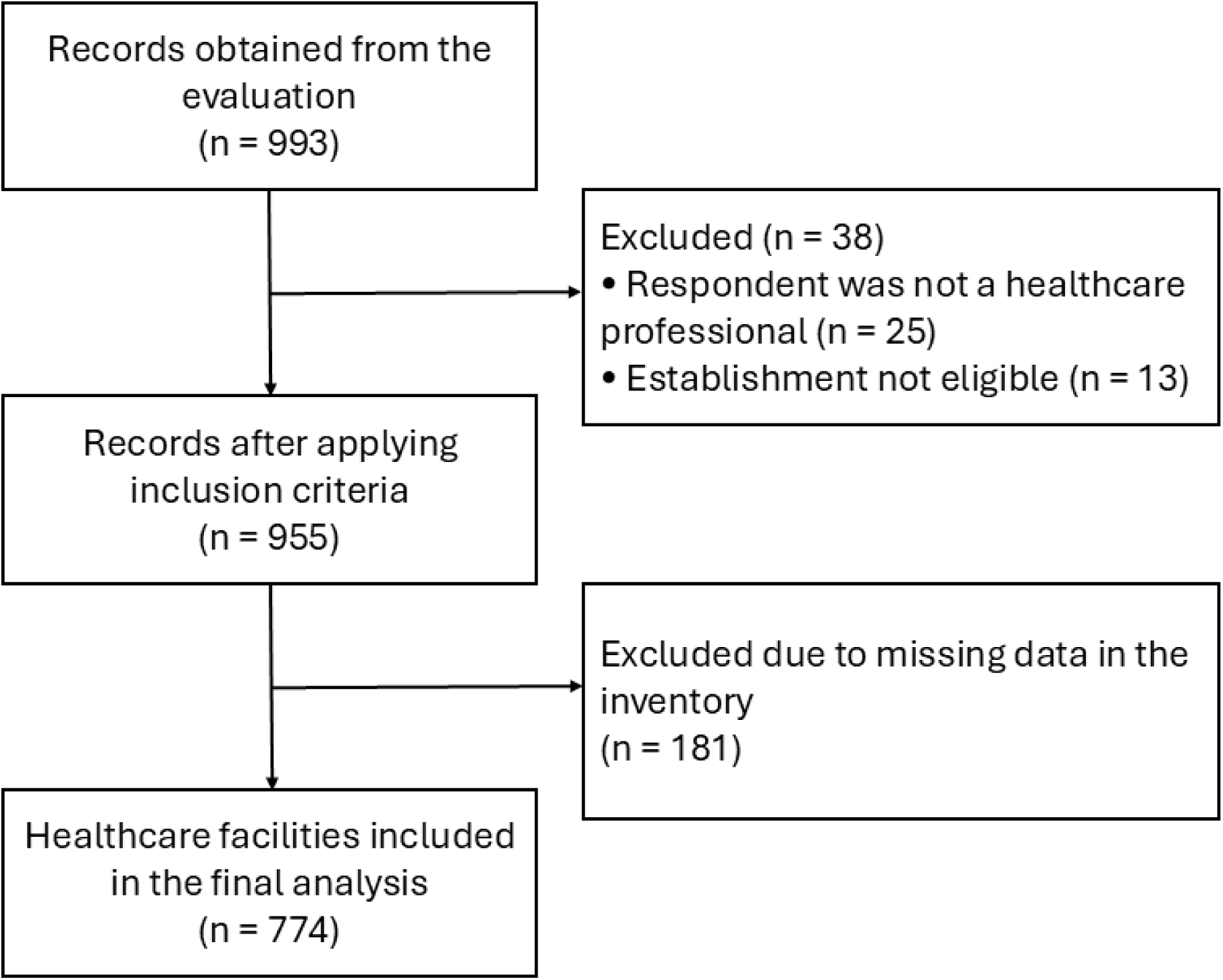
Sample selection flowchart.

**Table 2.**
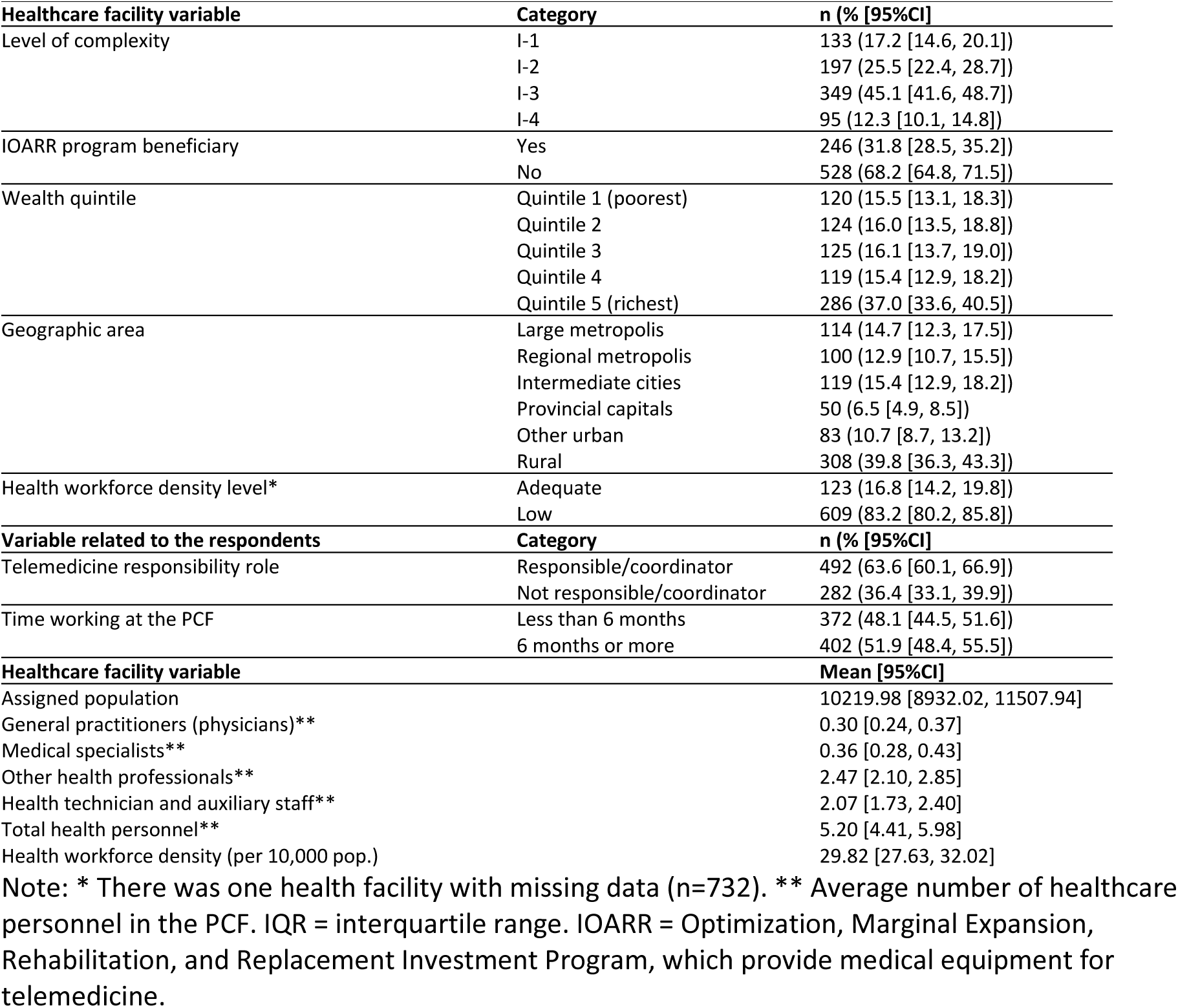
Demographic characteristics of the healthcare facility and healthcare personnel who responded (n = 774).

### Exploratory Factor Analysis

The total sample (n = 774) was randomly split into two independent datasets. EFA was conducted using data from 394 PCFs, whereas CFA was performed using data from the remaining 380 facilities.

We examined unidimensional solutions and found that the Organizational Readiness dimension explained 40% of the variance. However, we removed item 6 and 17 from this dimension because their factor loadings did not meet the expected threshold (see Supplementary Material 2). For the remaining dimensions, all items showed factor loadings greater than 0.40. In addition, we report the factor loadings for the Processes scale (variance explained: 73%), Digital Environment (41%), Human Resources (56%), and Regulatory Issues (57%) in Supplementary Materials 3–6, respectively.

### Confirmatory Factor Analysis

Based on the EFA results, a CFA was conducted using the remaining half of the sample. The CFA showed that none of the models derived from the EFA achieved optimal goodness-of-fit indices. Therefore, an iterative correlated-error analysis was performed by removing items involved in high correlated residuals (>0.30) until adequate goodness-of-fit indices were obtained across all models (Table 3). The full iterative item-removal process is presented in Supplementary Material 7. Then, the final CFA was developed in the remaining 39 items.

**Table 3.** Confirmatory factor analysis fit indices for the stable factor structures identified in the exploratory stage (n = 380).

| Domain | Retained items | X<sup>2</sup> | df | CFI | TLI | RMSEA (90% CI) | SRMR | α | ω |
| --- | --- | --- | --- | --- | --- | --- | --- | --- | --- |
| Organizational Readiness | 15 | 226.8 | 90 | 0.959 | 0.952 | 0.063 (0.053-0.074) | 0.057 | 0.86 | 0.86 |
| Processes | 5 | 11.2 | 5 | 0.998 | 0.997 | 0.057 (0.005-0.103) | 0.016 | 0.87 | 0.88 |
| Digital Environment | 8 | 49.5 | 20 | 0.967 | 0.953 | 0.062 (0.041-0.084) | 0.053 | 0.75 | 0.76 |
| Human Resources | 5 | 6.5 | 5 | 0.999 | 0.997 | 0.028 (0.000-0.081) | 0.026 | 0.78 | 0.79 |
| Regulatory Issues | 6 | 23.4 | 9 | 0.989 | 0.982 | 0.065 (0.033-0.098) | 0.037 | 0.79 | 0.79 |

Note: $\chi^2$ = chi-square test statistic; df = degrees of freedom; CFI = Comparative Fit Index; TLI = Tucker–Lewis Index; RMSEA = Root Mean Square Error of Approximation; CI = confidence interval; SRMR = Standardized Root Mean Square Residual; $\alpha$ = Cronbach’s alpha; $\omega$ = McDonald’s omega.

### Internal Consistency Analysis

The point estimates of reliability are presented in Table 3. In all cases, the five scales showed optimal levels of internal consistency for both the α (>0.75) and ω (>0.75) coefficients.

### Measurement Invariance Analysis

A measurement invariance analysis was conducted according to facility complexity level (I-1/I-2 vs. I-3/I-4) and time working at the PCF (less than 6 months vs. 6 months or more). These grouping variables were dichotomized to meet the requirements of the invariance analysis. The results indicated that the five domains were invariant across both grouping variables, with ΔCFI values below 0.010 in all comparisons. Therefore, the five domains can be considered measurement invariant, supporting meaningful comparisons between these groups. Moreover, all models showed adequate goodness-of-fit indices for CFI, TLI, RMSEA, and SRMR (see Table 4).

**Table 4.** Measurement invariance across health facility category, telemedicine responsibility role, and time working at the PCF (n = 774).

| Group | Domain | Invariance level | chisq | df | CFI | TLI | RMSEA (90% CI) | SRMR | ΔCFI | ΔRMSEA |
| --- | --- | --- | --- | --- | --- | --- | --- | --- | --- | --- |
| Health facility category<br>(I-1/I-2 vs I-3/I-4) | Digital Environment | Configural | 116.8 | 40 | 0.954 | 0.935 | 0.071 (0.056-0.086) | 0.057 |  |  |
|  |  | Thresholds | 134.4 | 48 | 0.948 | 0.939 | 0.068 (0.055-0.082) | 0.057 | -0.006 | -0.002 |
|  |  | Metric | 142.5 | 55 | 0.947 | 0.946 | 0.064 (0.051-0.077) | 0.059 | -0.001 | -0.004 |
|  | Human Resources | Configural | 38.8 | 10 | 0.987 | 0.974 | 0.086 (0.059-0.116) | 0.036 |  |  |
|  |  | Thresholds | 46.4 | 15 | 0.986 | 0.981 | 0.074 (0.050-0.098) | 0.036 | -0.001 | -0.013 |
|  |  | Metric | 67.3 | 19 | 0.978 | 0.977 | 0.081 (0.061-0.103) | 0.042 | -0.008 | 0.008 |
|  | Organizational Readiness | Configural | 518.1 | 180 | 0.952 | 0.944 | 0.070 (0.063-0.077) | 0.061 |  |  |
|  |  | Thresholds | 536.6 | 195 | 0.952 | 0.948 | 0.067 (0.061-0.074) | 0.061 | -0.001 | -0.002 |
|  |  | Metric | 514.4 | 209 | 0.957 | 0.956 | 0.062 (0.055-0.068) | 0.061 | 0.005 | -0.006 |
|  | Processes | Configural | 34.0 | 10 | 0.998 | 0.996 | 0.079 (0.051-0.109) | 0.017 |  |  |
|  |  | Thresholds | 39.4 | 15 | 0.998 | 0.997 | 0.065 (0.041-0.090) | 0.017 | 0.000 | -0.014 |
|  |  | Metric | 42.1 | 19 | 0.998 | 0.998 | 0.056 (0.033-0.079) | 0.017 | 0.000 | -0.009 |
|  | Regulatory Issues | Configural | 38.0 | 18 | 0.994 | 0.991 | 0.054 (0.030-0.078) | 0.033 |  |  |
|  |  | Thresholds | 41.4 | 24 | 0.995 | 0.994 | 0.043 (0.019-0.065) | 0.033 | 0.001 | -0.010 |
|  |  | Metric | 37.4 | 29 | 0.998 | 0.998 | 0.028 (0.000-0.050) | 0.034 | 0.003 | -0.016 |
| Time working at the PCF<br>(Less than 6 month vs 6 months or more) | Digital Environment | Configural | 121.4 | 40 | 0.957 | 0.939 | 0.073 (0.058-0.088) | 0.057 |  |  |
|  |  | Thresholds | 135.6 | 48 | 0.953 | 0.946 | 0.069 (0.055-0.083) | 0.057 | -0.003 | -0.004 |
|  |  | Metric | 158.1 | 55 | 0.945 | 0.944 | 0.070 (0.057-0.083) | 0.060 | -0.008 | 0.001 |
|  | Human Resources | Configural | 28.4 | 10 | 0.992 | 0.983 | 0.069 (0.040-0.100) | 0.034 |  |  |
|  |  | Thresholds | 34.8 | 15 | 0.991 | 0.988 | 0.059 (0.033-0.084) | 0.034 | -0.001 | -0.011 |
|  |  | Metric | 38.2 | 19 | 0.991 | 0.991 | 0.051 (0.027-0.075) | 0.036 | 0.000 | -0.007 |
|  | Organizational Readiness | Configural | 533.5 | 180 | 0.951 | 0.943 | 0.071 (0.064-0.078) | 0.060 |  |  |
|  |  | Thresholds | 558.9 | 195 | 0.950 | 0.946 | 0.070 (0.063-0.076) | 0.060 | -0.001 | -0.002 |
|  |  | Metric | 523.7 | 209 | 0.957 | 0.956 | 0.063 (0.056-0.069) | 0.061 | 0.007 | -0.007 |
|  | Processes | Configural | 23.1 | 10 | 0.999 | 0.998 | 0.058 (0.027-0.090) | 0.015 |  |  |
|  |  | Thresholds | 37.7 | 15 | 0.998 | 0.997 | 0.063 (0.038-0.088) | 0.015 | -0.001 | 0.004 |
|  |  | Metric | 37.0 | 19 | 0.998 | 0.998 | 0.050 (0.025-0.073) | 0.015 | 0.000 | -0.013 |
|  | Regulatory Issues | Configural | 42.0 | 18 | 0.994 | 0.989 | 0.059 (0.036-0.082) | 0.032 |  |  |
|  |  | Thresholds | 45.0 | 24 | 0.994 | 0.993 | 0.048 (0.025-0.069) | 0.032 | 0.001 | -0.011 |
|  |  | Metric | 51.8 | 29 | 0.994 | 0.994 | 0.045 (0.024-0.065) | 0.034 | -0.001 | -0.003 |

Note: $\chi^2$ = chi-square test statistic; df = degrees of freedom; CFI = Comparative Fit Index; TLI = Tucker–Lewis Index; RMSEA = Root Mean Square Error of Approximation; CI = confidence interval; SRMR = Standardized Root Mean Square Residual; ΔCFI = change in CFI; $\Delta$ RMSEA = change in RMSEA. The configural model was used as the reference model; therefore, ΔCFI and $\Delta$ RMSEA are not reported for this level. Invariance was considered supported when ΔCFI ≤ 0.010 and $\Delta$ RMSEA ≤ 0.015 between successive models.

### Percentile Values and Criterion Validity

In Table 5, normative values for the five domains are presented across percentile-based categories. The complete questionnaire, instructions, and response options are provided in Supplementary Material 8. We found that all five domains showed positive but small correlations with proportion of telemedicine use (p<0.001). These findings suggest that, although each domain is directly associated with telemedicine use, its relationship with these operational outcomes is modest or limited (see Table 5).

**Table 5.** Normative Values Based on Percentiles and Correlations with Telemedicine Utilization Indicators for the Five Domains (n = 773).

| Statistic |  | Digital Environment | Human Resources | Organizational Readiness | Processes | Regulatory Issues |
| --- | --- | --- | --- | --- | --- | --- |
| Percentile | p1 | 8 | 5 | 17 | 5 | 6 |
|  | p5 | 10 | 5 | 24 | 5 | 7 |
|  | p10 | 11 | 6 | 26 | 5 | 8 |
|  | p15 | 12 | 7 | 28 | 5 | 9 |
|  | p25 | 14 | 8 | 31 | 5 | 11 |
|  | p50 (Median) | 17 | 10 | 37 | 9 | 14 |
|  | p75 | 21 | 13 | 43 | 14 | 17 |
|  | p85 | 23 | 15 | 46 | 15 | 18 |
|  | p90 | 24 | 15 | 48 | 16 | 19 |
|  | p95 | 26 | 17 | 51 | 19 | 20 |
|  | p99 | 29 | 20 | 57 | 20 | 24 |
| Descriptive | M | 17.43 | 10.37 | 37.11 | 9.87 | 13.86 |
|  | SD | 5.08 | 3.58 | 8.50 | 4.50 | 4.30 |
| Levels | Low | 8-14 | 5-8 | 15-31 | 5-5 | 6-11 |
|  | Medium | 15-20 | 9-12 | 32-42 | 6-13 | 12-16 |
|  | High | 21-32 | 13-20 | 43-60 | 14-20 | 17-24 |
| Spearman correlation | Proportion of telemedicine use | 0.21* | 0.19* | 0.22* | 0.15* | 0.17* |

Note: We constructed percentiles as low (minimum value to the 25th percentile), medium (26th to 74th percentile), and high (75th percentile to the maximum value). The first outcome was defined as the proportion of telemedicine use; the second outcome was defined as total telemedicine consultations divided by the assigned population. \*All correlations were statistically significant at $p < .001$ . The sample size was 773 because we had one missing data point for the variable "proportion of telemedicine use."

## Discussion

### Main findings

Our study adapted and validated the Telemedicine Readiness Inventory at the Facility Level. The adapted instrument demonstrated a stable factor structure, adequate internal consistency, evidence of measurement invariance, and a positive association between the overall score and facility-level telemedicine use, suggesting preliminary criterion validity. These findings indicate that the instrument has adequate psychometric properties, providing evidence of validity and reliability and supporting its use in primary care settings. Although the PAHO/IDB source tool was originally framed as a maturity assessment tool, the adapted version is better understood as a telemedicine readiness instrument because it assesses institutional conditions for implementation rather than a prescriptive pathway for progressing across maturity levels [27,40,41].

Furthermore, our findings suggest that a higher level of institutional telemedicine readiness, as assessed across the five domains, is associated with greater telemedicine adoption in practice. Facilities with higher readiness scores tended to report greater telemedicine use. Although the magnitude of this association was small, it was positive and statistically significant. This finding is consistent with previous studies indicating that telehealth readiness can improve the success of telemedicine implementation [42]. However, access to telemedicine remains unequal worldwide. Approximately 75% of high-income countries report the use of telemedicine [43,44], whereas only around 10% of healthcare facilities in low-income settings offer telemedicine services [44].

### Dimensionality

Our study identified the five domains previously proposed by the PAHO/IDB scale (Organizational Readiness, Processes, Digital Environment, Human Resources, and Regulatory Issues) [27]. However, it provides the first psychometric and quantitative evaluation of the evidence supporting these five domains. The original WHO-IDB instrument did not report evidence of validity or reliability [27]. We excluded the Expertise domain included in the original version of the instrument because it could not be adequately operationalized and, based on the recommendation of the study’s expert panel, it was removed from the final model. Although this may be considered a limitation, the construct could still be partially assessed, and we consider that excluding this complementary domain is unlikely to compromise the overall evaluation of telemedicine readiness.

Previous studies applied the original PAHO/IDB scale with all six dimensions and reported descriptive findings on telemedicine maturity levels in Nigeria, Uruguay, and Peru, without providing psychometric evidence to support the validity or reliability of these results [45–47]. Moreover, in settings with different health system characteristics, the original dimensional structure may not be replicable, reducing confidence in the certainty and interpretability of previous findings. One example of these interpretability concerns is that all identified studies using the original PAHO/IDB tool reported overall scores, means, or percentages based on a general instrument score. However, to generate scores that meaningfully represent a latent dimension, the corresponding items should exhibit similar factor loadings or, at a minimum, sufficiently strong loadings to support such analyses. In our study, several items showed factor loadings below commonly accepted thresholds. Therefore, it is possible that the same limitation affected previous applications of the instrument.

Compared with earlier readiness tools, which were often supported primarily by expert judgment or content validity [48–50], our study adds evidence on factor structure, reliability, measurement invariance, and criterion-related validity in a large sample of PCFs. Furthermore, we did not identify any other telemedicine readiness instrument that has undergone a psychometric evaluation or formal validation study. Psychometric studies have been conducted for related constructs, such as the four-dimensional Digital Hospital Evaluation Scale and the six-dimensional Questionnaire to Measure Digital Maturity of General Practitioner Practices [51,52]. Although their factorial structures were derived empirically rather than from locally grounded frameworks or local quantitative studies [53]. While empirically derived models may fit the data well, they may also be less closely aligned with local implementation realities, and this can limit the broader generalizability of their findings across contexts.

### Measurement Invariance

Our analyses found that the instrument’s measurement properties were equivalent across different facility subgroups (scalar and metric invariance), meaning that the item-dimension relationships held consistently across groups. Our study analyzed two relevant sources of potential differences in the data: an institutional difference related to facility complexity, defined by level of care, and a potential source of bias related to whether the respondent knew the information used to assess the PCF, defined by their length of work within the facility. In our study, the instrument was invariant across both variables. This indicates that both groups understood the construct equivalently for each domain, allowing the use of the same norms and cut-off points, as well as comparisons between groups.

Differences in training, institutional capacity, and technological readiness within the health system can affect how institutions develop capacities to provide services such as telemedicine [54,55]. However, our study suggests that the same instrument can validly and reliably assess these potential differences within primary care health systems. In practical terms, our invariant structure implies that differences in readiness scores between both groups can be interpreted equivalently. Without invariance, one could not be sure if a higher readiness score in one group truly reflected greater readiness or was simply an error of how questions function [38]. Thus, our findings provide robust support that the adapted tool measures telemedicine readiness equivalently in diverse healthcare settings, allowing its use for comparisons and benchmarking across groups.

### Criterion Validation

Because there is no universal gold standard for telemedicine readiness, observed telemedicine use is best interpreted as a pragmatic external criterion that reflects adoption, but not necessarily quality, safety, continuity, appropriateness, or equity [1,8]. Our study found small positive associations between the five readiness domains and telemedicine use. This finding may reflect the fact that readiness functions as an enabling condition rather than a direct determinant of service volume. Telemedicine use is also influenced by multiple factors, including patient demand, connectivity, digital literacy, incentives, organizational culture, clinical appropriateness, and the regulatory environment [6,11,12].

### Implications

TRI-F has important implications for digital health policy and practice. It can serve as a diagnostic tool for health systems to identify facility-level gaps that impede telemedicine scale-up. As WHO’s Global Strategy on Digital Health emphasizes, telemedicine is a key component for achieving universal health coverage by improving access, continuity, and efficiency of care [56]. Our results support calls to integrate telehealth readiness into broader quality frameworks. For example, US accreditation programs now include telehealth standards: NCQA’s Virtual Care Accreditation provides organizational quality benchmarks in structural components of virtual care, enabling institutions to identify program gaps and drive improvement.

Readiness assessment should be positioned as an initial component of a broader telemedicine implementation architecture that includes needs assessment, planning, change management, service implementation, performance monitoring, and continuous improvement [1,8,57]. Future studies should build on the validated TRI-F to develop a prescriptive telemedicine maturity model that links domains and levels to concrete improvement actions, responsible actors, resources, monitoring indicators, and measures of quality, safety, equity, and sustainability [22,40,41].

In practice, percentile-based norms may help facilities compare their readiness scores with those of similar facilities in the local context, but they should not be interpreted as universal cutoffs for low, medium, or high readiness [21,57]. In addition, our adapted tool can support public-sector decision-making by identifying facility-level gaps, prioritizing technical assistance, guiding investments, and monitoring readiness over time. However, it should not be used as a rigid certification of high-quality telemedicine delivery [1,57].

### Strengths and limitations

Our study has three main strengths. First, we evaluated a large number of primary care health facilities, providing sufficient statistical power for the various analyses conducted. Second, to our knowledge, this is the first study to perform a rigorous quantitative evaluation of the instrument properties of the PAHO/IDB inventory. Third, our tool operationalizes the response options, reducing reliance on subjective perceptions of item fulfillment that characterized the original version.

However, our study also has some limitations. First, we used self-reported data from a single informant per facility, which may introduce measurement bias by incomplete knowledge, and social desirability effects. Nevertheless, we found measurement invariance between professionals with more than 6 months of work experience at the facility and those with 5 months or less wich might suggest small or non significant effects of incomplete knowledge of PCF. Therefore, although these factors could introduce bias, we consider its potential influence to be small. Second, the sample came from a national network related to telehealth, so the results should not be generalized directly to all public primary care facilities. Third, the cross-sectional design prevents causal inference between readiness and telemedicine use. Fourth, in the criterion validity analysis, the telemedicine use indicators did not capture quality, safety, continuity, clinical appropriateness, equity, patient experience, or patient-side barriers. Therefore, the relationship with real-world telemedicine performance in health facilities may have been assessed only partially. Fifth, the instrument mainly measures supply-side readiness through the five identified domains: Organizational Readiness, Processes, Digital Environment, Human Resources, and Regulatory Issues. However, it does not fully cover system-level factors such as financing, clinical training, or patient-side barriers, and therefore may partially capture the construct. Nevertheless, the original study proposed a clear theoretical framework for telemedicine readiness [27], supporting the consistency of the construct evaluation.

## Conclusions

This study provides robust psychometric evidence for the TRI-F, demonstrating satisfactory structural validity, internal consistency, and measurement invariance across key institutional and respondent subgroups. The positive, albeit modest, associations between readiness scores and telemedicine utilization highlight that readiness is an enabling condition, but not a sufficient determinant of sustained, equitable, and high-quality telemedicine delivery. Accordingly, the tool should be used primarily as a diagnostic and monitoring tool to identify facility-level gaps, guide investment decisions, and prioritize technical assistance, rather than as a certification of telemedicine performance. Future implementation efforts and policy initiatives should embed telemedicine readiness assessment within broader strategies that also evaluate service quality, clinical appropriateness, patient experience, equity, safety, and long-term sustainability.

## Declarations

## Acknowledgments

The authors thank the Ministry of Health of Peru for facilitating access to the requested information used in this study. We also acknowledge the Directorate General of Telehealth, Referral and Emergency Services (DIGTEL) for its role in coordinating the telemedicine readiness assessment and supporting the availability of institutional information. The contents of this manuscript, including the analyses, interpretation, and conclusions, are solely the responsibility of the authors and do not necessarily represent the official position of the Ministry of Health or DIGTEL.

The authors also thank Jesse Sanchez-Vargas and Mariel Portal for their early input during the initial development of this work.

## Funding

This study was self-funded.

### Availability of data and materials

The database and code are available at: https://doi.org/10.6084/m9.figshare.32514540

### Consent to publication

Not applicable.

## Competing interests

The authors do not report any conflict of interest when conducting the study, analyzing the data, or writing the manuscript.

### Declaration of generative AI and AI-assisted technologies in the writing process

We used DeepL to translate specific sections of the manuscript and Grammarly to improve the wording of certain sections. All authors reviewed and approved the final version of the manuscript.

### Authors’ contributions

Stefan Escobar-Agreda: Conceptualization, Methodology, Validation, Investigation, Data Curation, Writing - Original Draft.

David Villarreal-Zegarra: Conceptualization, Methodology, Validation, Formal analysis, Investigation, Data Curation, Writing - Original Draft, Visualization.

Yscenia Paredes-Gonzales: Validation, Investigation, Writing - Review & Editing.

C. Mahony Reategui-Rivera: Methodology, Validation, Investigation, Writing - Original Draft.

Leonardo Rojas-Mezarina: Conceptualization, Validation, Investigation, Writing - Review & Editing, Supervision.

---

**Supplementary material 1.**
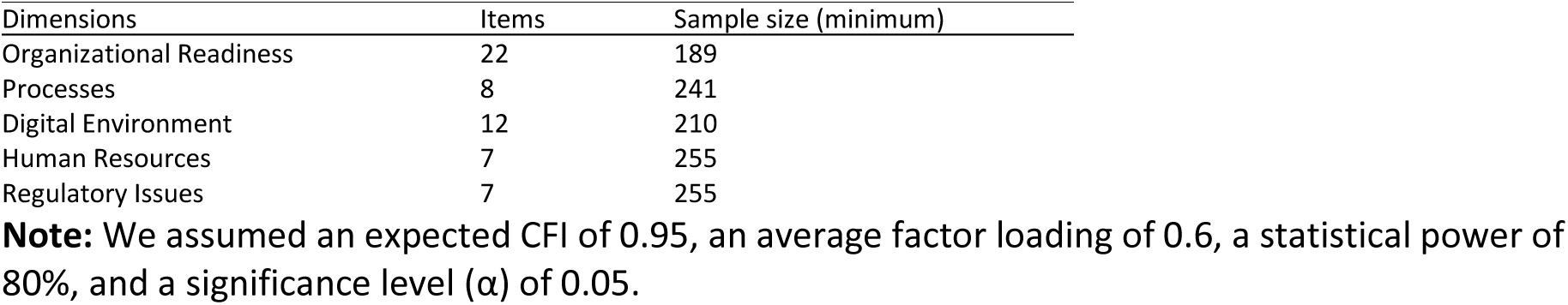
Calculation of sample size by confirmatory factorial analysis.

**Supplementary material 2.**
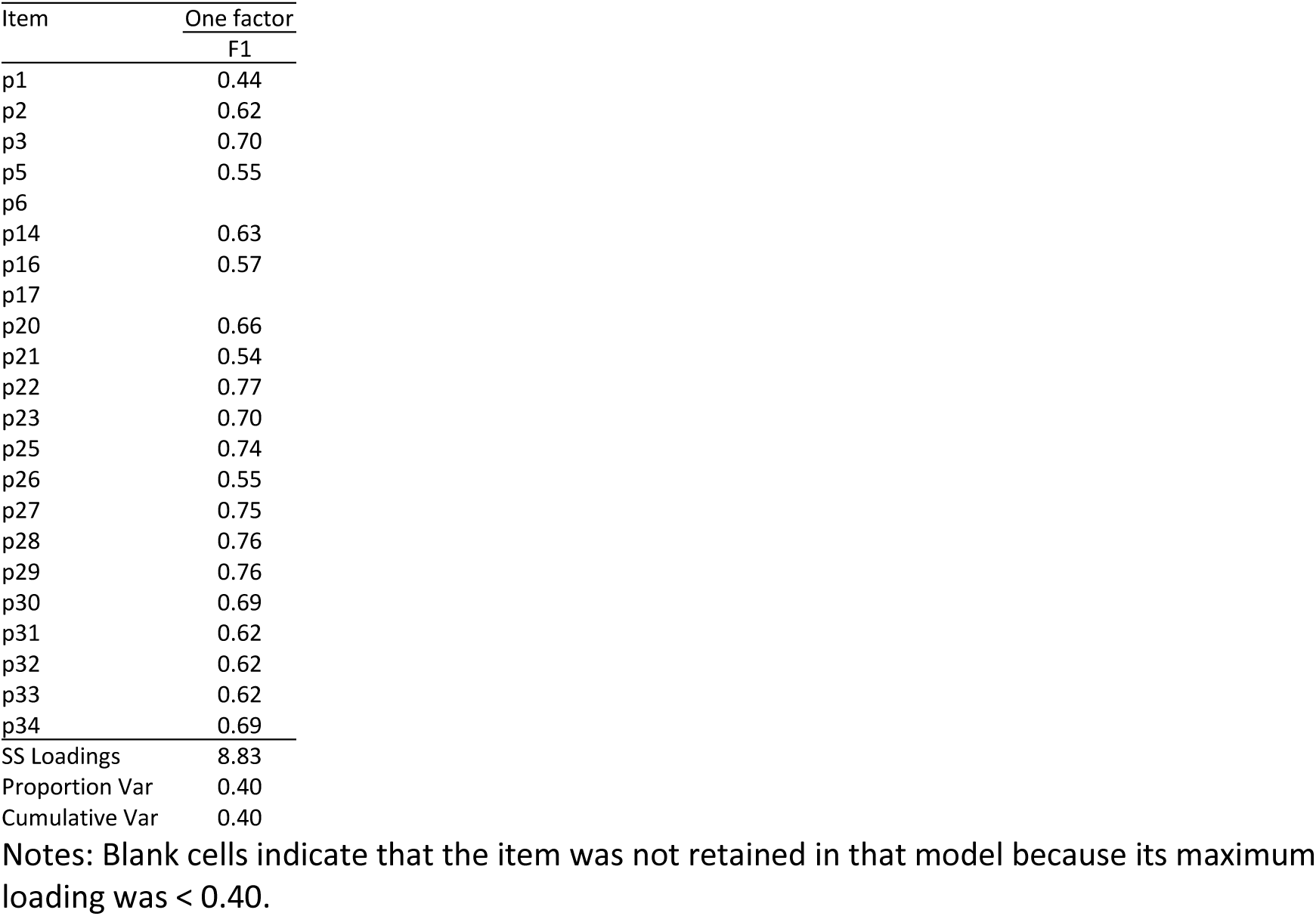
Organizational Readiness (n=394).

**Supplementary material 3.**
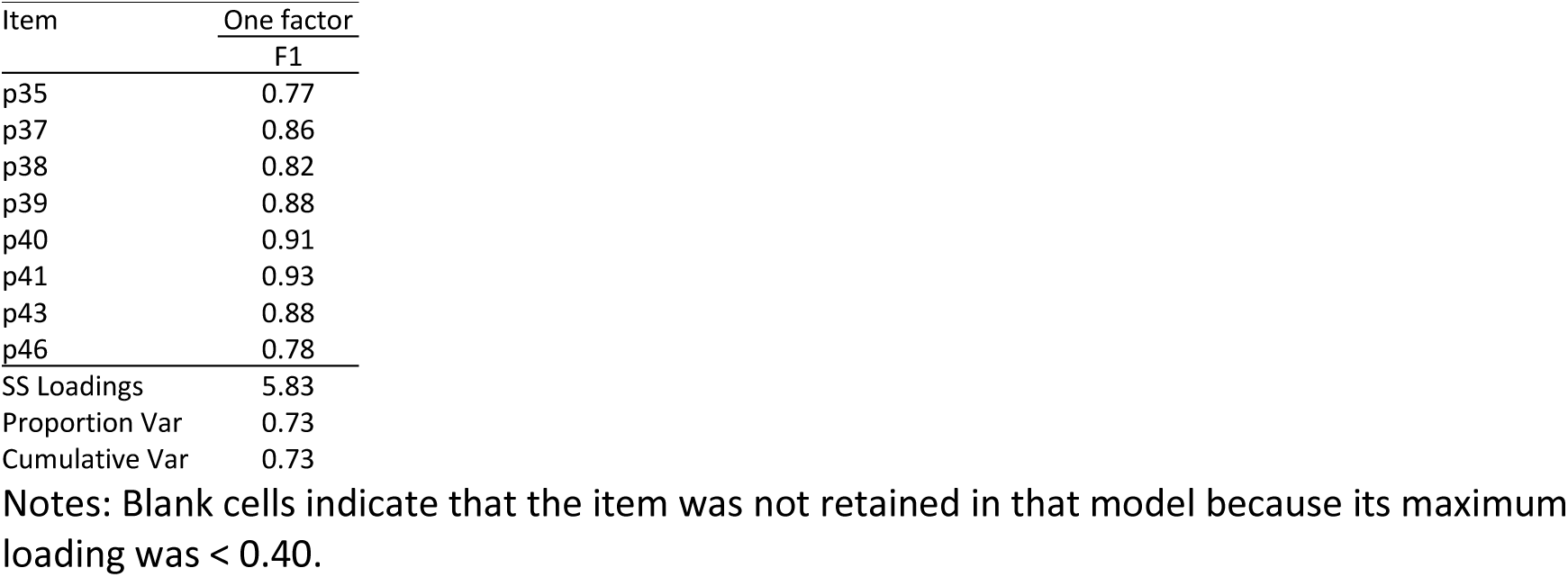
Processes (n=394).

**Supplementary material 4.**
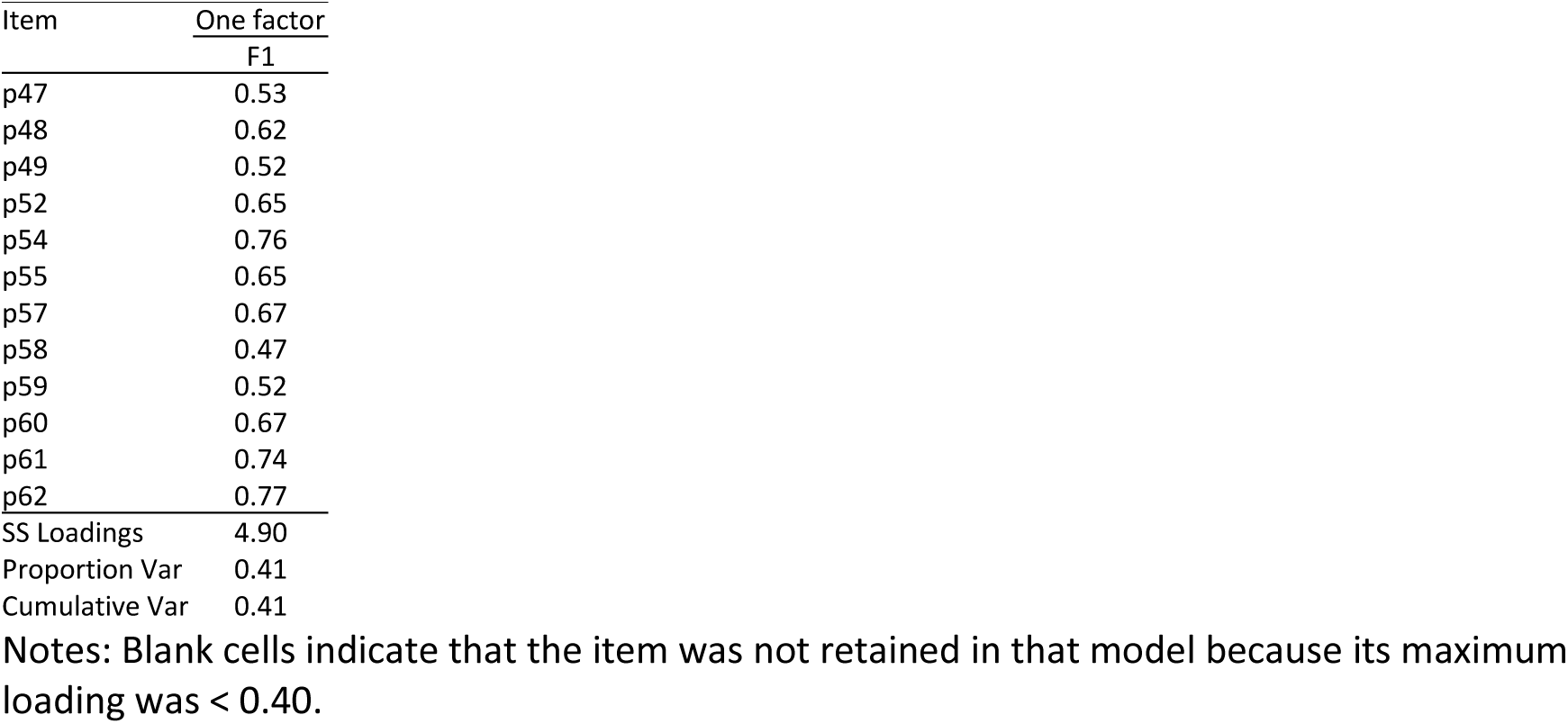
Digital Environment (n=394).

**Supplementary material 5.**
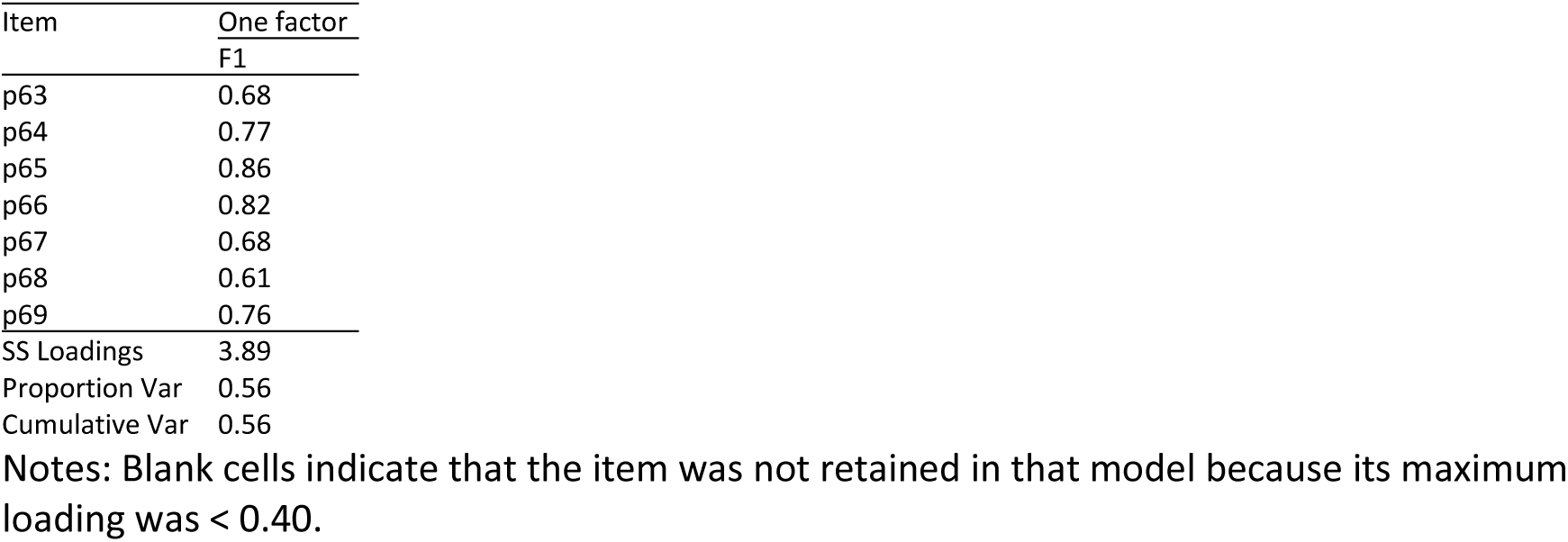
Human Resources (n=394).

**Supplementary material 6.**
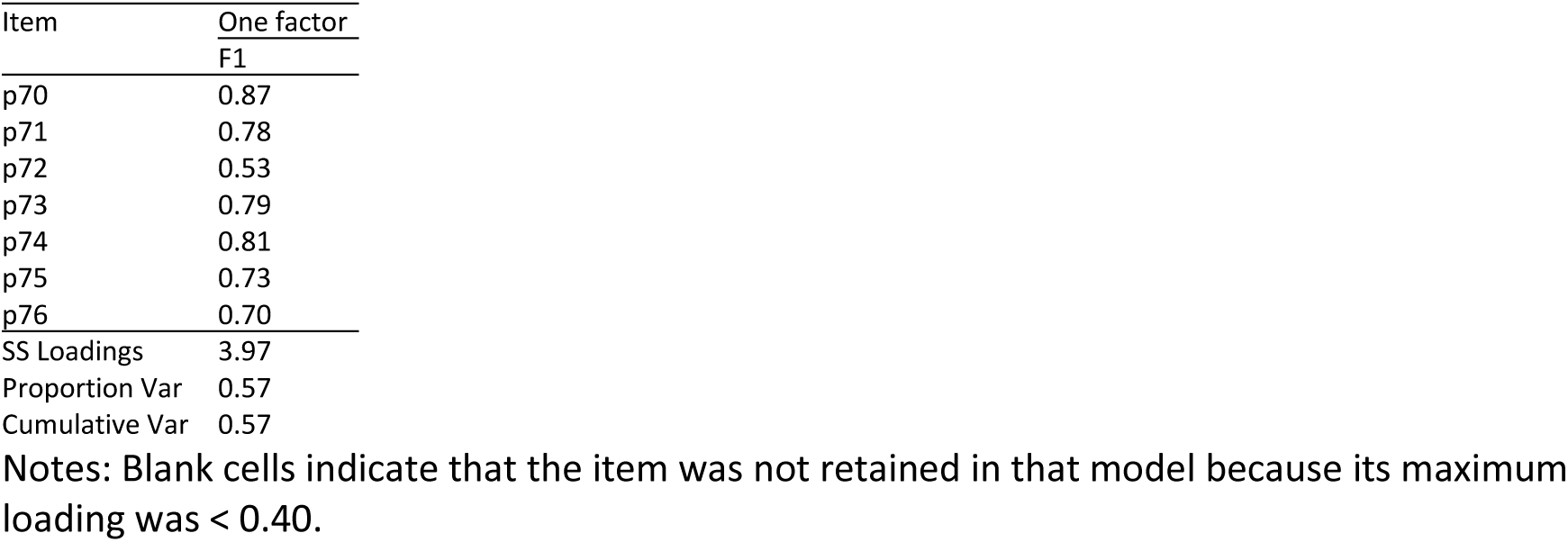
Regulatory Issues (n=394).

**Supplementary material 7.**
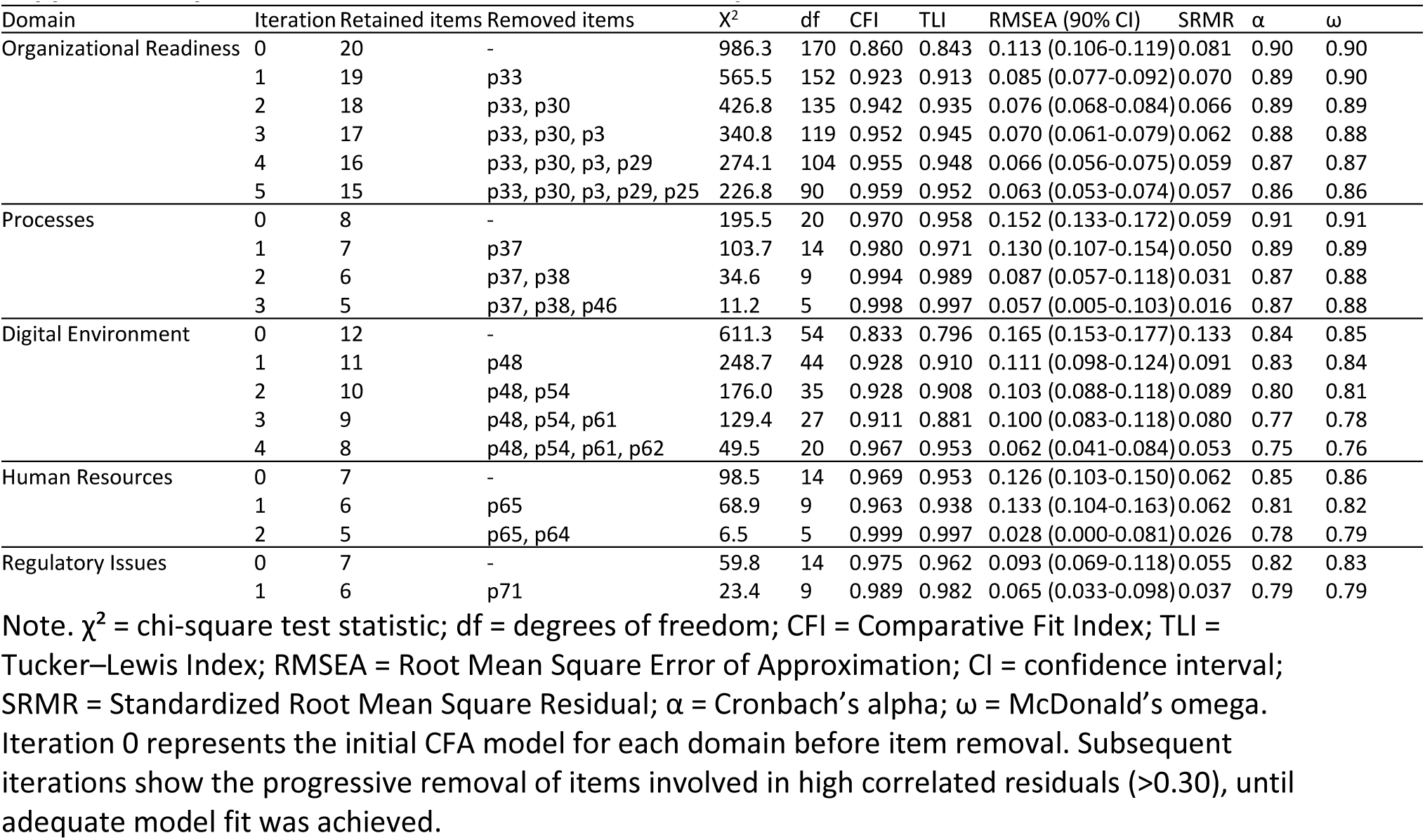
Iterative CFA item-removal process based on correlated residuals.

**Supplementary material 8.**
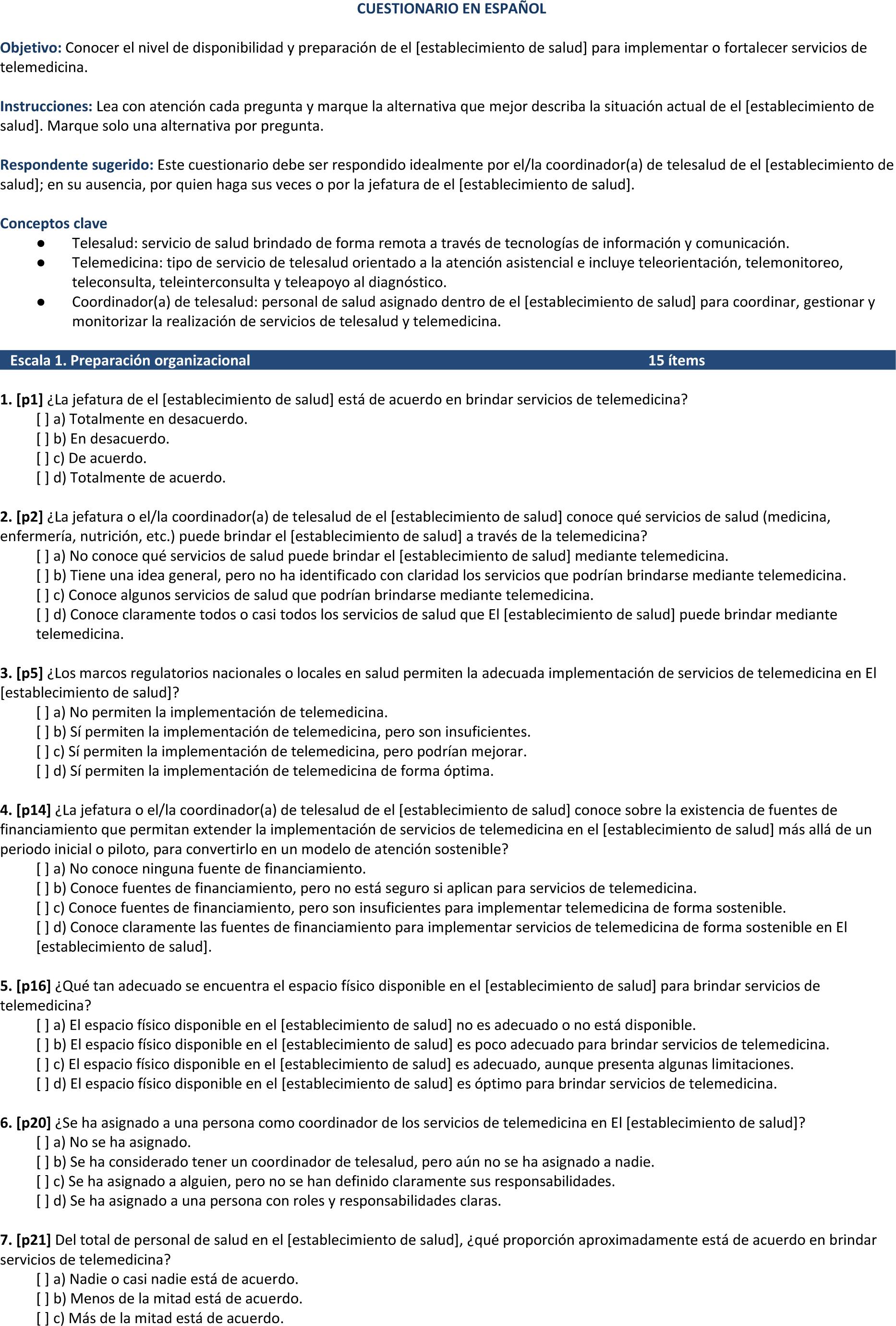
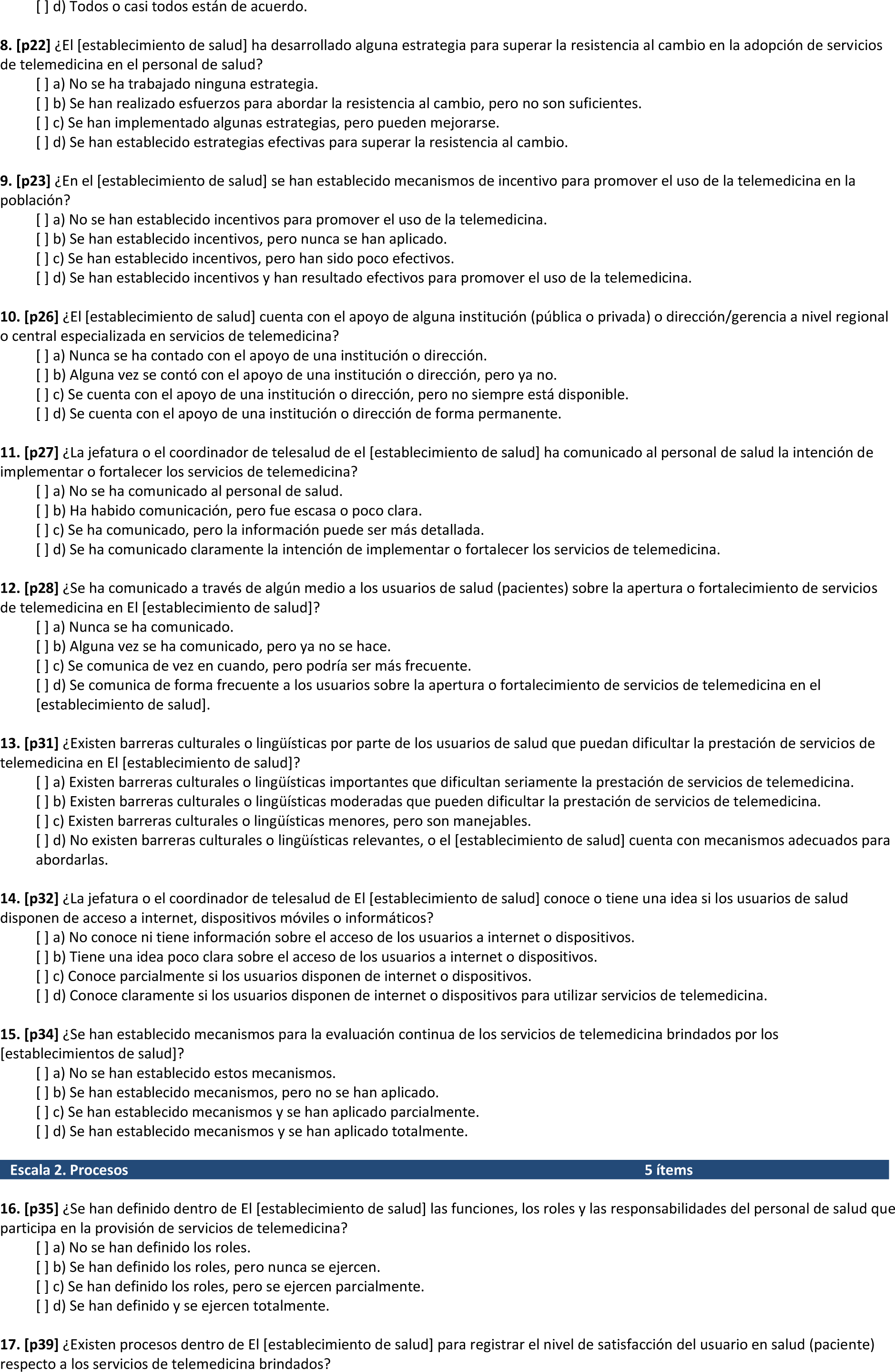
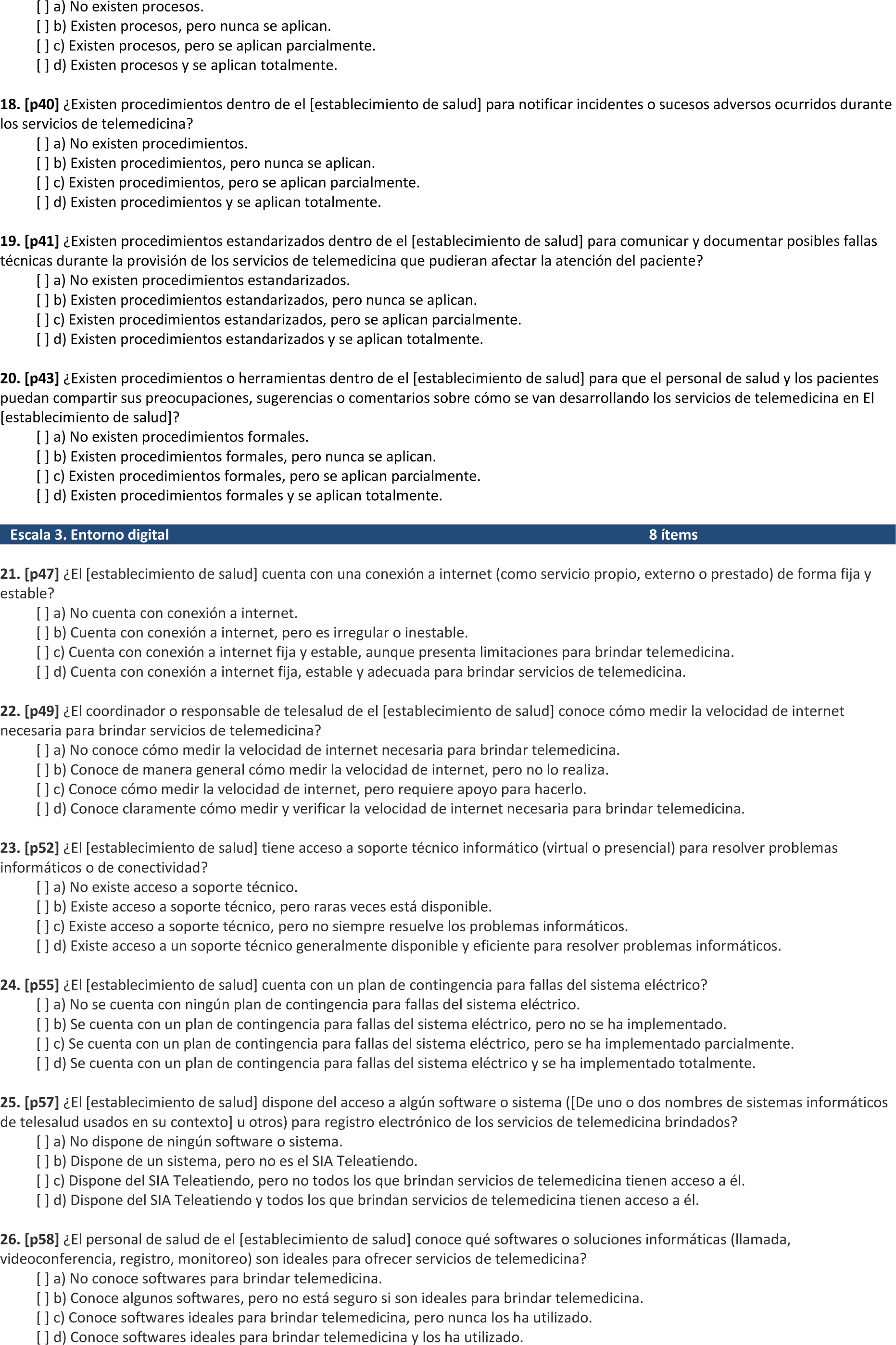
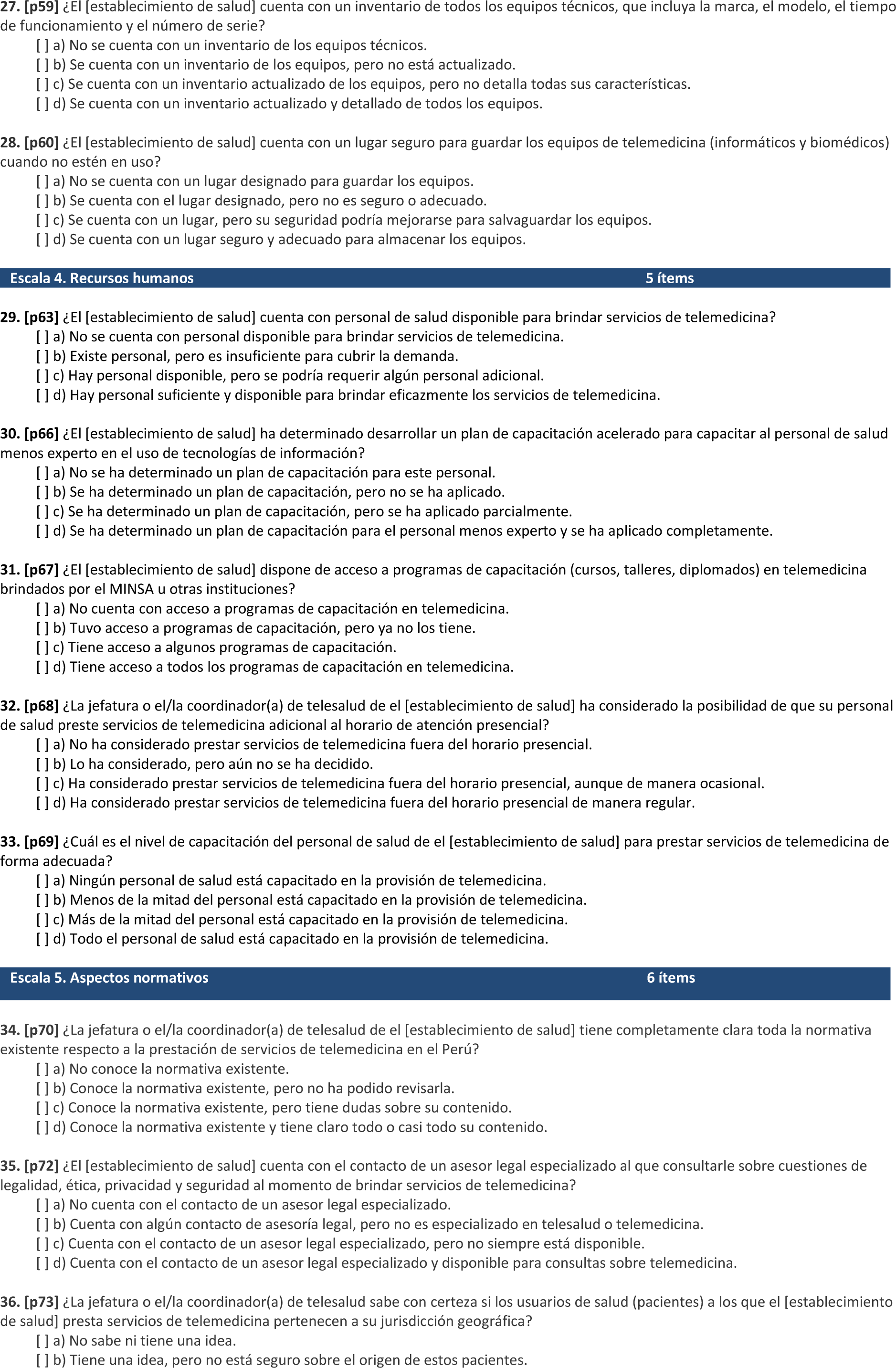
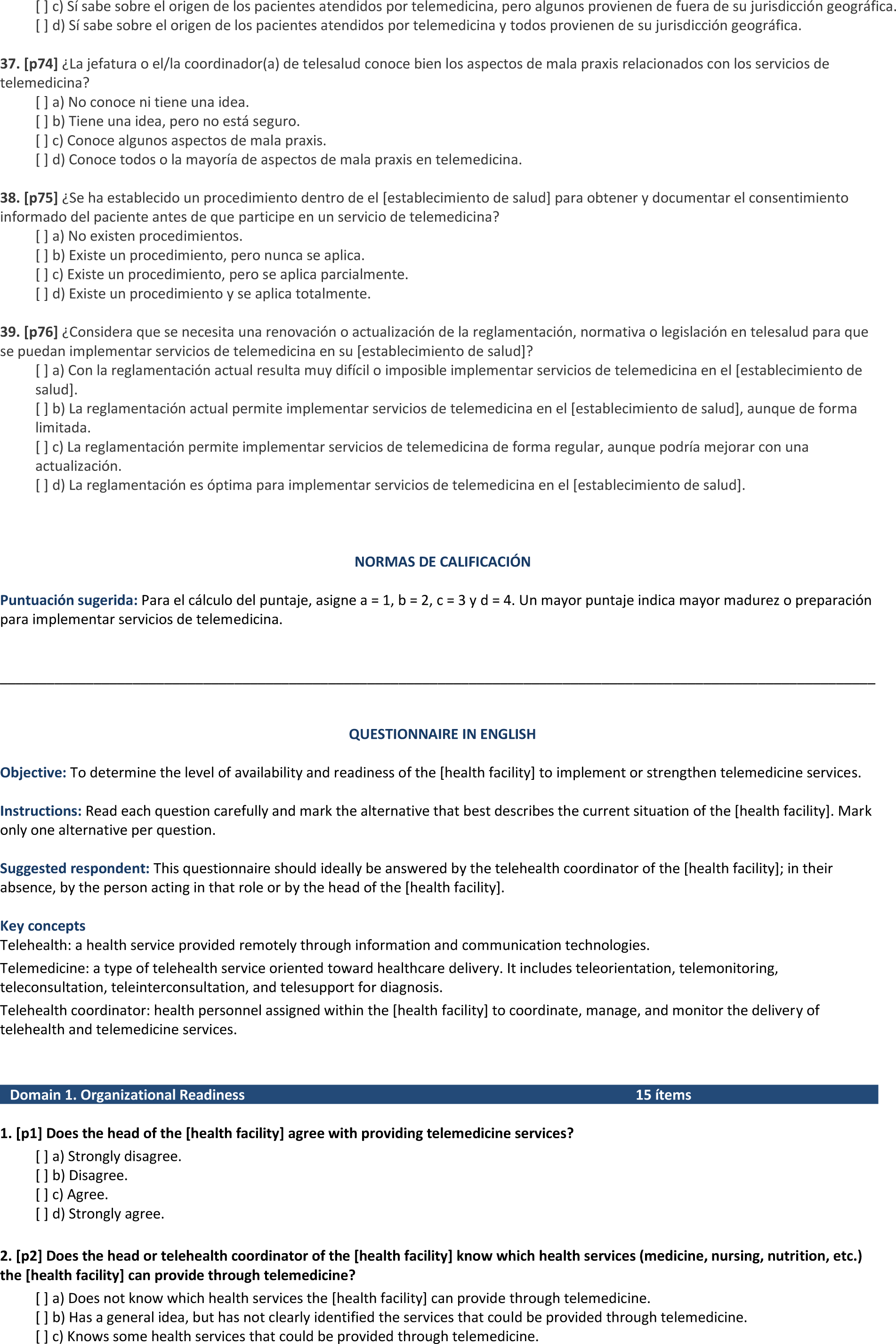
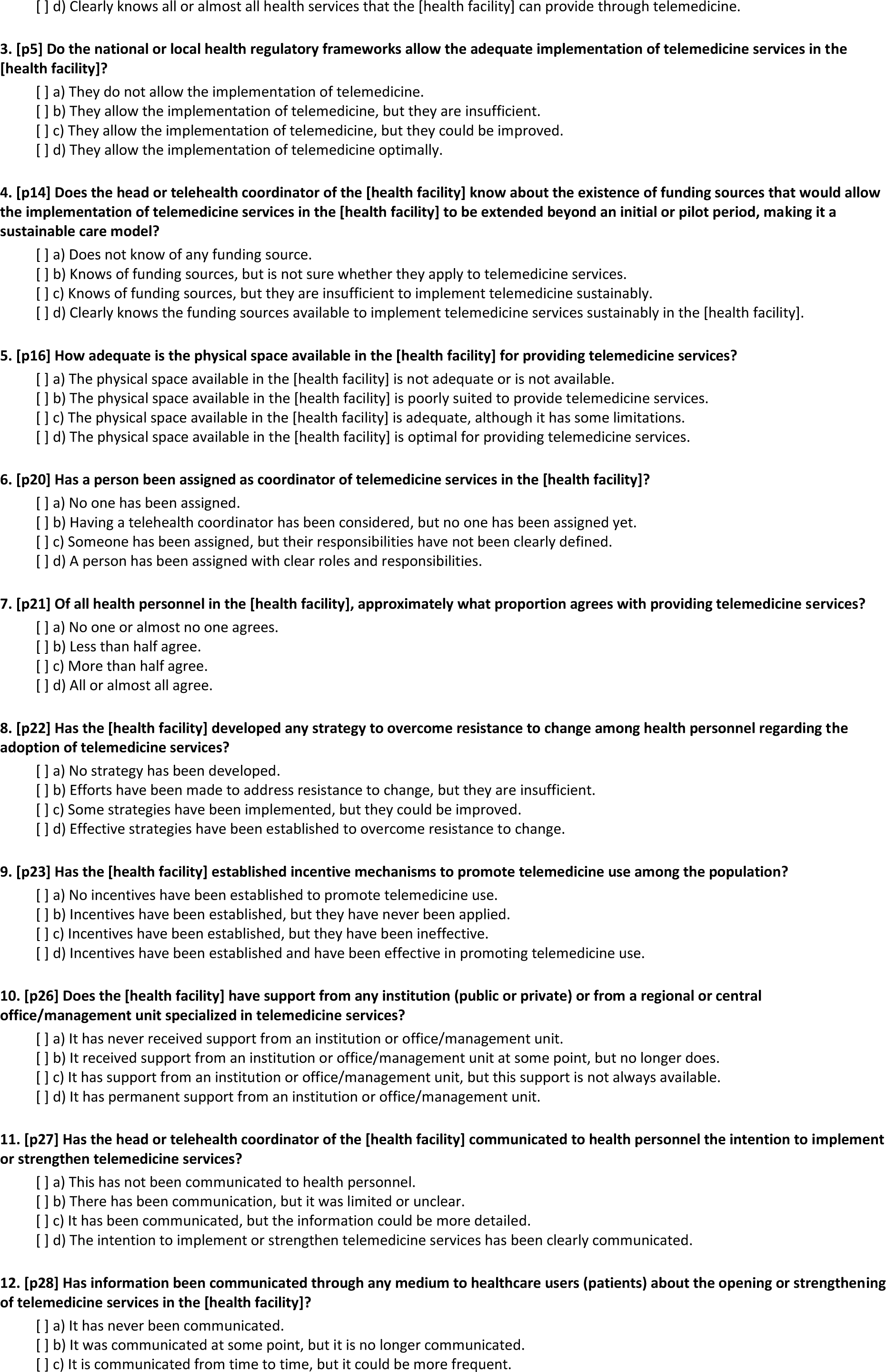
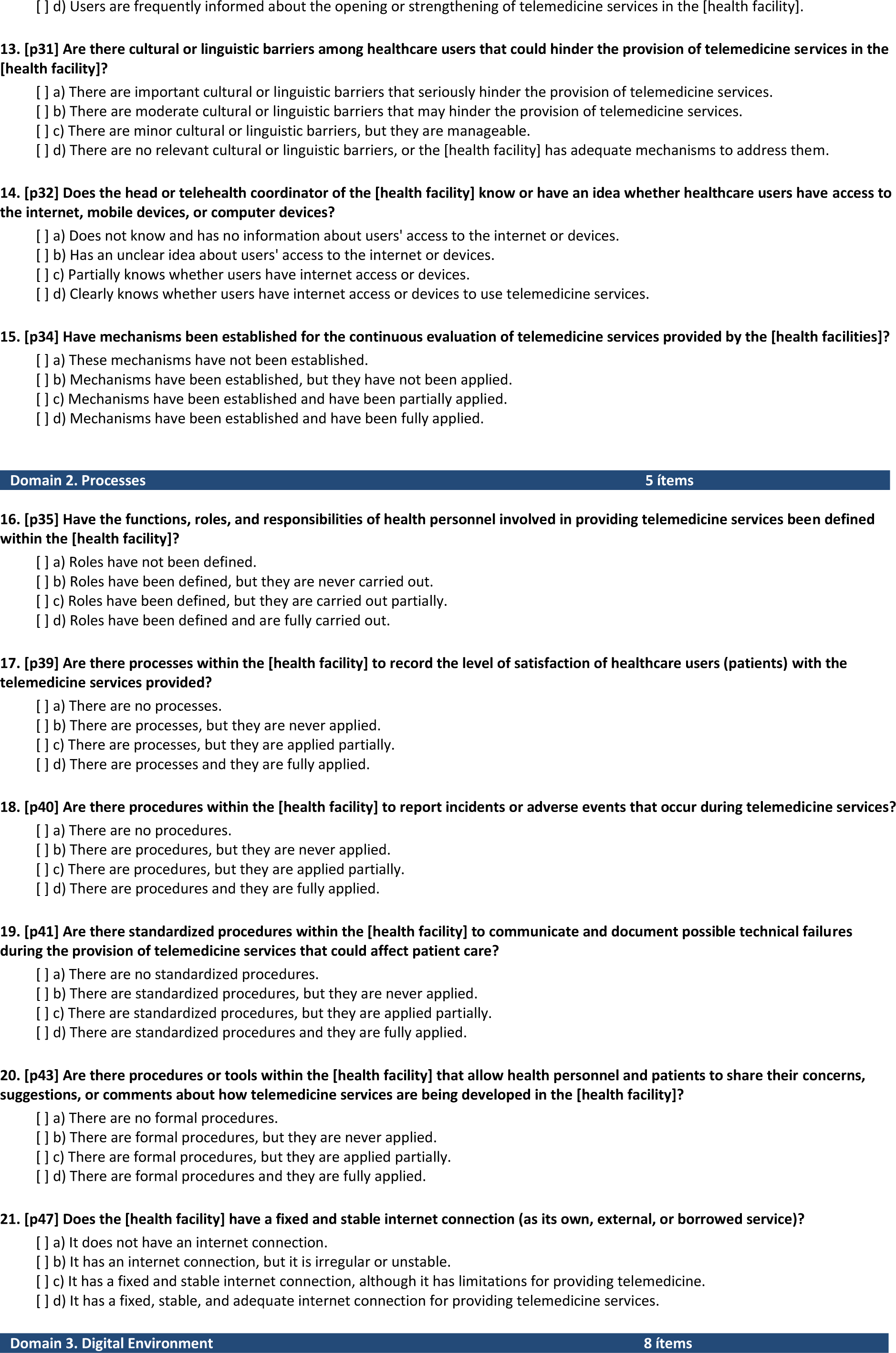
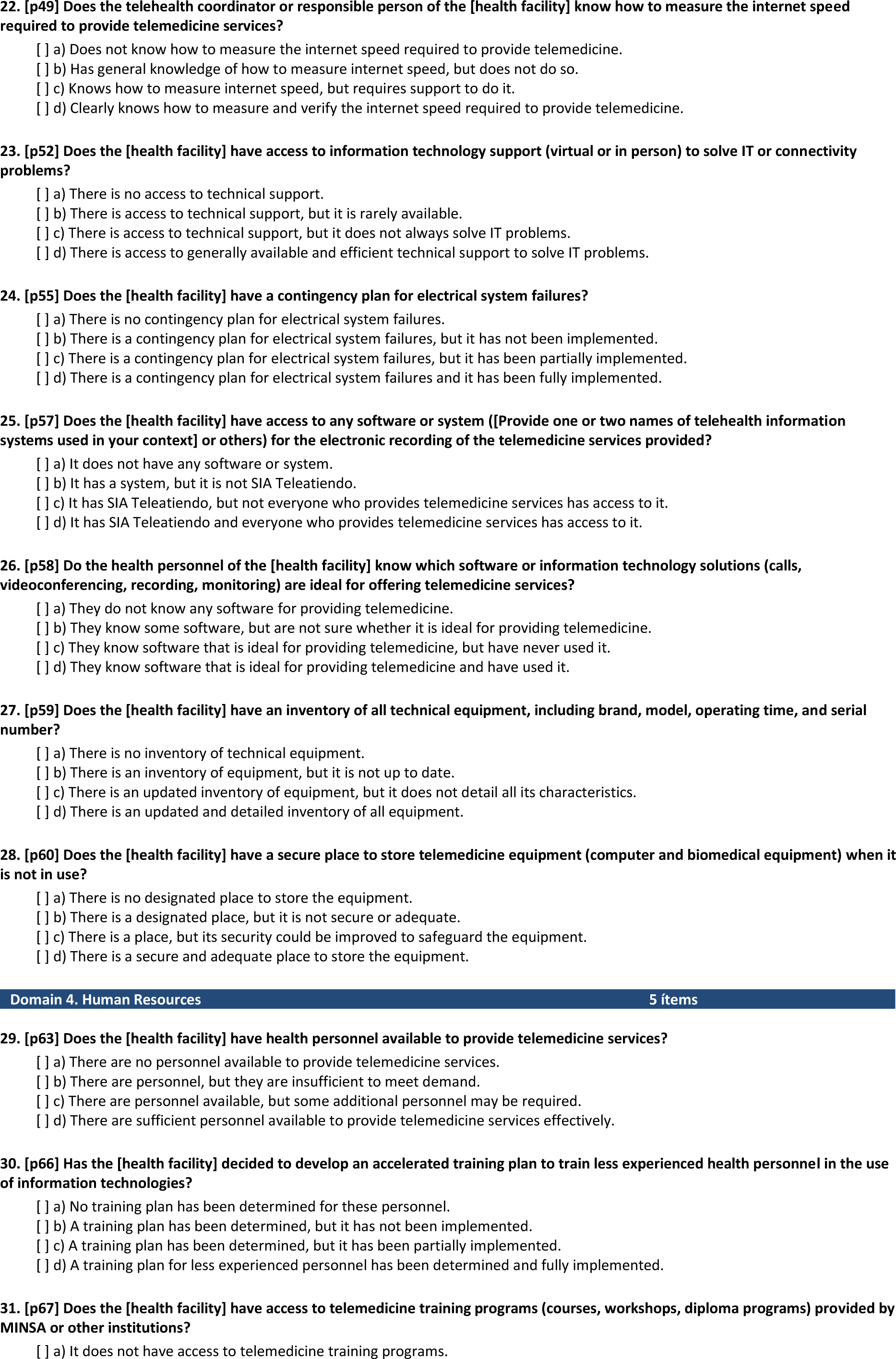
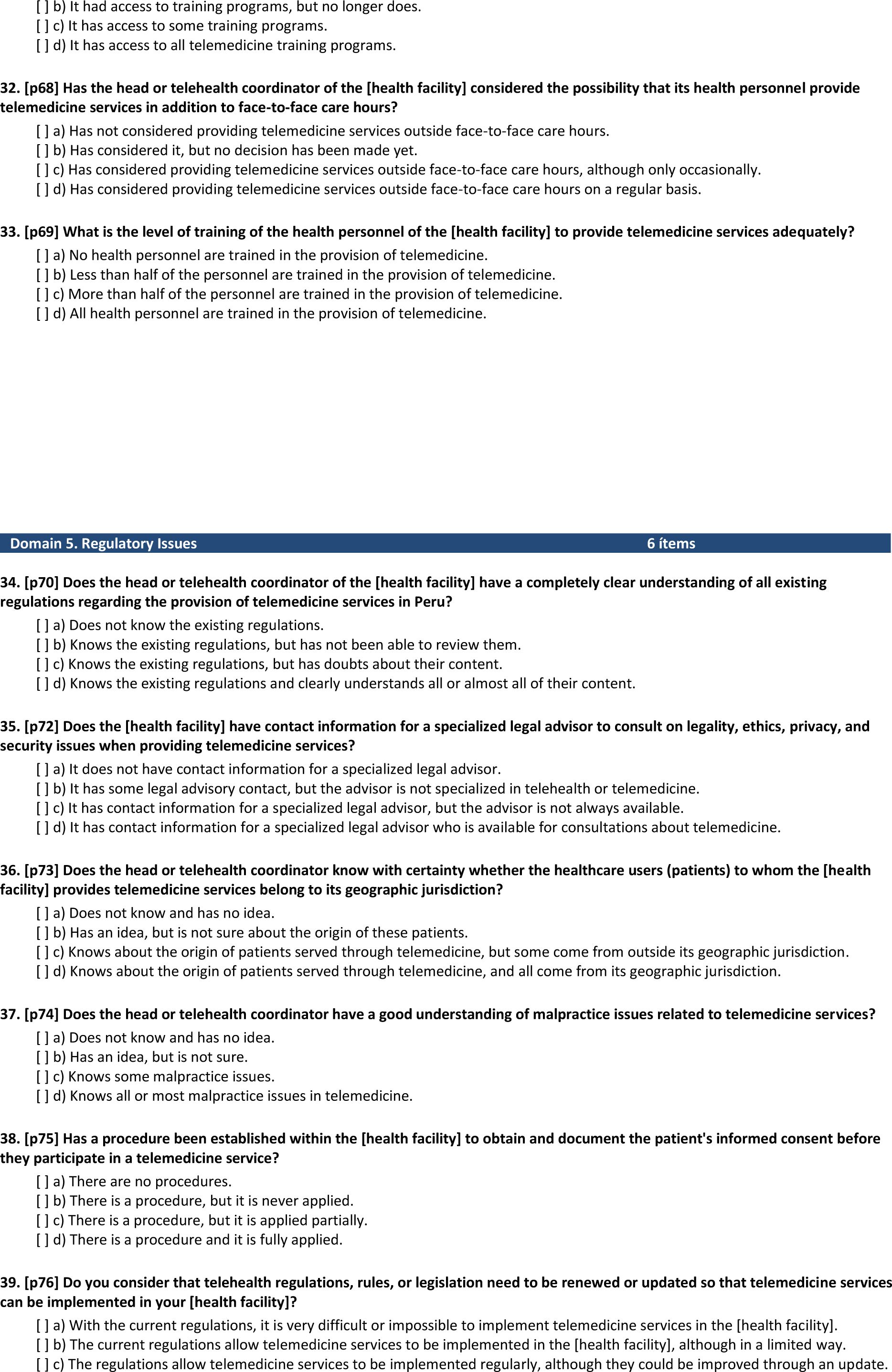
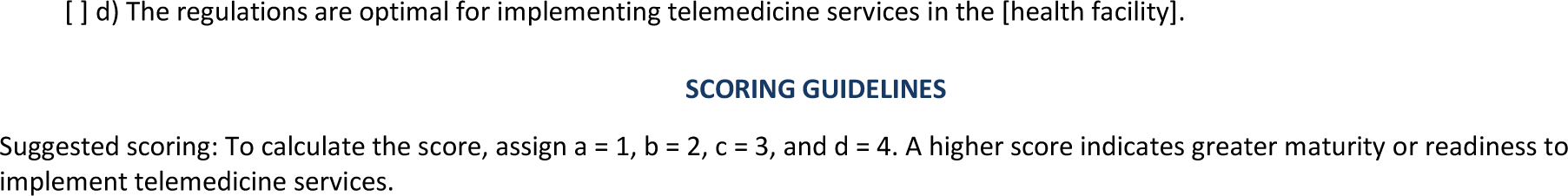
Telemedicine Readiness Tool, in Spanish and English.

